# Multicomponent Blood Pressure Control Program in Medicare Beneficiaries with Hypertension Primarily of Hispanic Origin in a Large Health System in South Florida

**DOI:** 10.1101/2025.05.29.25328595

**Authors:** Paulo H. M. Chaves, Maureen DeSoria, Leslye Roque, Rafael J. Mas

## Abstract

**Background:** Health care systems play a key role in hypertension control in large populations, which remains challenging. This study assessed: (a) blood pressure (BP) control rate changes pre-vs. post-implementation of a hypertension control program in the understudied Hispanic population in South Florida; and (b) compared annual BP control rate agreement rates calculated using time-average BP (AVG-BP) vs. BP from most recent visit (LAST-BP).

**Methods:** Leon Medical Centers (LMC), a major integrated healthcare services provider to Medicare and dual eligible patients in Miami-Dade, implemented a multicomponent hypertension control program in 2011. Electronic health records (EHRs) data from patients aged 65-89 years with hypertension were analyzed. Longitudinal analysis involved calculation of age- and diabetes-standardized annual BP control rates, and random effects modeling. Systolic<140 mmHg and diastolic<90 mmHg defined controlled BP. Kappa statistic was calculated.

**Results:** From 2008 (n=4,710) to 2018 (n=21,540), mean age ± SD increased (73.4±5.3 to 77.1±6.1). Proportion of Hispanics remained constant (>98%). BP increased substantially, similarly in women and men post-program implementation; e.g., in women, standardized control rates (AVG-BP) increased ranged from 68.9-73.3% in 2008-2010 to 88.8-92.3% in 2013-2018. Improvement observed using LAST-BP was analogous, though latter absolute rates were systematically lower. Kappa between AVG-BP- and LAST-BP-derived rates in 2018 was 0.296.

**Conclusions:** Program implementation was associated with a meaningful, sustained increase in BP control up to high levels. Agreement between AVG-BP and LAST-BP control rates was only fair. Whether longitudinal EHRs longitudinal BP data-derived metrics may offer added-value in health services-related assessments remains to be determined.

## INTRODUCTION

Uncontrolled hypertension is a major causal and potentially modifiable risk factor for increased morbidity and mortality ^1–3^. Despite the fact that efficacious intervention approaches for blood pressure (BP) control in adults with hypertension exist ^4–8^, BP control rates in large populations remain disappointingly low ^9–11^. For example, recent analysis of the US National Health and Nutrition Examination Survey (NHANES) data from 2015-2016, documented an overall hypertension control (BP<140/90 mm Hg) rate of less than 50% in non**-**institutionalized adults aged 60 years and older, the subset in which hypertension prevalence is highest ^12^.

Extensive research effort to date has revealed that health care systems may play a key role in enhancing BP control in large populations via implementation of a team care-based approach addressing multiple health care delivery components ^3,6,7,13^. However, data remain scarce on the effectiveness of large scale, multi-component hypertension control programs in minority segments of the population in which the burden of uncontrolled hypertension is high, including the understudied population of older adults of Hispanic origin in South Florida. Research in this latter large segment is particularly relevant, as BP control rates in Hispanics, analogously to as in Blacks, lower than that in non-Hispanic Whites have been reported ^12^. In the Hispanic Community Health Study/Study of Latinos, for instance, the percent of controlled BP (<140/90 mm Hg) among participants 65 years and older with health insurance was only 41% in men, and 38% in women, which was lower than in NHANES non-Hispanic white participants^14^. In another study that involved cross-sectional analysis of electronic health record (EHR) data from 2012-2016 gathered as part of a large scale collaborative clinical research network in Florida, documented uncontrolled BP control rates (<140/90 mm Hg) of 46.6% (n=74,340) in patients aged 65-74 years old, and 53.2% (n=72,875) in patients aged 75 years and older were reported ^15^.

Methodologically, evaluation of the effectiveness of health systems in controlling BP in patients with hypertension in a specific year traditionally involves the use of snapshot BP obtained at a single time-point, traditionally the most recent BP reading available in a year, for calculation of annual BP control rates ^16,17^. Although a snapshot BP measure offer a valid approach that optimizes clinical feasibility, important caveats should be acknowledged, including the fact that BP snapshots are prone to random measurement error that can contribute to BP status misclassification and underestimation of adverse outcome risk ^18^. Additionally, snapshot BP at a single time point is inherently weaker than time-average BP is terms of capturing the cumulative adverse impact of high BP levels on health, as indicated by observational data demonstrating that the average of multiple systolic BP measurements over time obtained in the past may be related independently and more strongly to the incident risk of future adverse outcomes than a current systolic BP snapshot ^19,20^. With the broad implementation of electronic health records (EHRs), the routine capturing of repeated BP measurements on the same patient over time has now become feasible, at least in large health systems. The extent to which inferences on BP control in health systems may vary depending on whether single-time point snapshot BP or within-person time-average BP is used to define BP control, an important preliminary step into research ultimately aimed at leveraging serial BP data in EHR for improving healthcare delivery to older patients with hypertension, remains to be evaluated.

The goals of this study were: (a) to test the hypothesis that the implementation of a primary care-based program by a major health care services provider to Medicare and dual eligible outpatients primarily of Hispanic origin in Miami-Dade, Florida, was associated with improvement in BP control in older women and men with hypertension; and (b) to compare agreement in BP control classification between control rates calculated based on within-person annual-average BP and those based on a single-time point snapshot BP.

## METHODS

### Study Design

This was a secondary longitudinal data analysis study. Data came from the Leon Medical Centers (LMC)’s EHR system. LMC is a major integrated healthcare services provider to Medicare and dual eligible patients in Miami-Dade County, Florida (FL). LMC patients are enrollees in the Medicare Advantage plan that works exclusively with LMC, the Leon Medical Centers Health Plans. In 2011, LMC implemented a large-scale, primary care setting-based quality improvement program for BP control in patients, who are primarily of Hispanic origin, with hypertension. This study initially considered all health system patients aged 65 years and older with an International Classification of Diseases (ICD)-based diagnosis for HTN receiving care in one of 7 LMC outpatient centers located in Miami Dade, FL. Additional eligibility criteria for each study year included continuous enrollment between January 1 and December 31, and having at least 2 BP measures recorded in EHRs from regular outpatient visits, being at least one in each half of the study year. We subsequently restricted analysis to those aged less than 90 years based on preliminary analysis related to age standardized analysis to ensure a balanced age range in all study years (below). Diagnosis of end-stage renal disease, and/or non-acute inpatient setting admission were not exclusion criteria in our study.

### LMC Hypertension Control Program

The LMC Hypertension Control Program was launched in 2011 as a system-wide, mandatory quality improvement initiative across all LMC outpatient medical centers. The program, which remains in operation today, is centered on a highly coordinated, multidisciplinary, team-based approach supported by a robust EHR system. The program delivers a comprehensive multicomponent intervention in the primary care setting.

The team-based care model includes primary care physicians (PCPs), registered nurses (RNs), clinical pharmacists, and medical assistants (MAs), working in close coordination. Under RN leadership, a dedicated care management unit – the Gold Standard Team (GST) – was created to oversee hypertension care and coordinate follow-up. System-wide hypertension control policies were internally disseminated, and protocols for standardized blood pressure (BP) measurement were established. Practical baseline training was provided to clinical staff, followed by biannual continuing education sessions to maintain competencies and incorporate clinical updates. BP readings were obtained by MAs at every primary care visit as part of routine vital sign assessments, and also at cardiology and urgent care encounters. Per protocol, if an initial BP reading obtained via an automated monitor was out of range, a flag was generated in the EHR to prompt a manual BP reading using an aneroid sphygmomanometer. If the manual reading remained elevated, the PCP was automatically notified to repeat the measurement at the start of the encounter and consider treatment modification.

Uncontrolled BP triggered a standing order for clinical staff to assess adherence and reinforce patient education. In cases where BP remained uncontrolled or treatment was modified, an automatic order was generated for a follow-up BP check. The GST received daily EHR reports identifying patients in the hypertension registry who required follow-up, and was responsible for scheduling follow-up checks every three weeks until control was achieved. The GST monitored medical records for subsequent BP readings; if BP remained uncontrolled, the team alerted the responsible PCP and arranged a follow-up BP appointment with the MA. Additionally, patients received a mandatory phone call explaining the reason for the follow-up visit. Cancellations and no-shows were actively tracked. The GST communicated regularly with PCPs via secure EHR messaging to share updates and address concerns identified through continuous surveillance.

Physicians and other members of the care delivery team were encouraged to follow evidence-based hypertension guidelines at the program’s inception^21^ (18) and integrate relevant updates as appropriate. PCP education, including continuing medical education (CME) activities, was regularly conducted. Periodic email reminders were sent by leadership to reinforce training opportunities. Physicians were given access to an open pharmacy formulary to allow individualized treatment plans, especially for older patients with complex conditions. Through an EHR-based dashboard, PCPs accessed real-time clinical information relevant to hypertension care, including laboratory and diagnostic results (e.g., renal function, electrolytes, diabetes status, ECG, echocardiograms). Monthly performance feedback was provided to physicians, and BP control rates within their patient panels were linked to quality performance evaluations and financial incentives.

Patient engagement was a core component of the program. Quarterly chronic disease management seminars were offered to enhance health literacy, with a focus on hypertension and related conditions such as diabetes. All patients on the hypertension registry were eligible, and invitations were issued via mail and PCP referral. Educational brochures in both English and Spanish were made available in medical center waiting areas. The LMC marketing department also published a triannual magazine featuring health articles – including on hypertension – which was mailed to patients’ homes.

Pharmacy services played a pivotal role in promoting and sustaining medication adherence. Adherence was actively monitored through an electronic hypertension registry and supported by several system-level strategies. These included waived Medicare Part D copayments, automatic refills (prior to CMS policy changes), home delivery of medications, pharmacist–patient consultations, and physician follow-up in cases of nonadherence. Following the discontinuation of automatic refill allowances by CMS, pharmacy staff proactively contacted patients due for refills. If a patient declined, an alert was sent via the EHR system to the patient’s PCP, prompting personalized follow-up. To improve accessibility and convenience, patients were offered the option to pick up prescriptions at on-site satellite pharmacies immediately after clinic visits or to receive medications via home delivery.

Recognizing transportation as a key social determinant of health, LMC integrated transportation into the program’s delivery infrastructure. Unlimited transportation to clinic appointments and medication delivery to patients’ homes were offered at no cost, a component often overlooked in previous hypertension control initiatives.

Finally, the EHR system was integral to the program’s implementation and maintenance. It enabled the creation of a hypertension clinical registry, facilitated real-time alerts, supported e-prescribing, allowed provider panel tracking via dashboards and patient summary reports, and enhanced clinical decision-making at the point of care.

### Annual Blood Pressure Control Rates

Two indicators of annual BP control status were calculated: one based on within-person time-average BP, referred to as *time-average BP* (AVG-BP), and the other based on snapshot BP obtained at a single time-point, chosen at BP available at the last visit in a selected year, referred to as *last BP* (LAST-BP).

#### Annual Hypertension Control Based on Time-Average BP

Within-person average systolic was calculated in each study year as follows. First, for each patient, we calculated the mean of all systolic BP readings separately for the first (between January 1 and June 30) and second (July 1 – December 31) half of each study year. If only one reading was available in a particular half, then that BP reading was used. Second, we calculated the average within-person systolic BP by adding the patient’s average systolic BP in the first semester to that in the second semester and dividing by 2. Patients without at least one BP reading in each semester period were excluded. Average within-person diastolic BP was calculated analogously. Finally, for each person, BP was considered controlled in each year if within-person average systolic BP was less than 140 mmHg, and within-person average diastolic BP was less than 90 mmHg, regardless of age or disease status. That threshold was chosen on the basis that it was consistent with that in clinical guidelines at the time the program was implemented ^21^, and with the threshold adopted by the Centers for Medicare & Medicaid Services (CMS) ^16^.

#### Annual Hypertension Control Based on Last BP

Consistent, though not identical, with the approach developed by the National Committee for Quality Assurance in its Healthcare Effectiveness Data and Information Set (HEDIS®), a widely used set of performance measures in the managed care industry ^17^, the systolic and diastolic BP reading at the last visit for each study year was used as the basis to classify BP control status^16^. BP was defined as controlled if systolic was less than 140 mmHg, and diastolic BP less than 90 mmHg. If multiple BP measurements occur on the same date or noted in the chart on the same date, use the lowest systolic and lowest diastolic BP reading

### Additional Data

Other data analyzed included: (a) demographic characteristics; i.e., age, gender, and ethnicity (a person was considered of Hispanic origin if they identified Spanish as their preferred language), and outpatient clinic center; and prevalent diabetes, assessed according to the International Classification of Diseases (ICD) – 9 or 10 classification.

### Statistical Analysis

Exploratory data analyses involved calculation of means ± standard deviations (SD) and proportions of univariate, unweighted distributions of selected variables in each study year. Patients included in study analysis included all those who met inclusion criteria in each study year, thus no sampling approach was used. This longitudinal study involved an open cohort, as patients could move in (e.g., new health plan enrollees) or out (e.g., withdrawals) of the health system at any time during study years. As the patient population became substantially larger over time, changes in the population structure were noted. For example, populations in years after program implementation were older and had higher diabetes prevalence than populations in years before program implementation. Because of those variations in population structures, weights were used to standardize the population structure in each study yea, so that estimates in changes in BP control rates were not due simply to differences in population structure. In this context, we calculated weighted BP control rates, along with corresponding 95% confidence intervals (CI) that adjusted for differences in the combined annual age distribution and diabetes prevalence over time. The population structure in 2010, the year that preceded program implementation, was chosen arbitrarily for standardization purposes. Random effects multivariate logistic regression models adjusted for age and prevalent diabetes were used to estimate the odds of controlled blood pressure (yes vs. no) during the intermediate (initial 2 years of program inception) and post-implementation phases (last 6 study years), as compared to the period before program implementation. Models included random subject-specific intercept to account for within-subject correlation of multiple BP control assessments over time. Data are presented as odds ratios adjusted for time-varying age group (<75 vs. 75-89 years) and prevalent diabetes status (yes vs. no), along with corresponding 95% confidence intervals. Unweight and weighted percent agreement calculated the overall agreement of the control rate based on time-average BP and the control rate based on last BP in classifying similarly subjects as having BP controlled or not in the last 5 study years. Cohen’s Kappa statistics, an indicator of degree of agreement beyond chance alone. Analyses were stratified by gender, given known differences in BP awareness, prevalence, and control between women and men. Stata, version 16 (StataCorp, College Station, TX) was used.

## RESULTS

### Patient Characteristics

The number of patients with hypertension data analyzed ranged from 4,710 in 2008 to 21,540 in 2018 (**Table 1**). In that period, mean age (from 73.4±5.3 to 77.1±6.1), and the prevalence of diabetes (from approximately 35% to 49%) increased gradually over time, while the proportion of women (range: 55-58% range) and that of patients of Hispanic origin (range: 97-99%) remained stable across study years.

**Table 1.** Selected Characteristics of Leon Medical Centers Patients with Hypertension Included in Analysis, Miami, Florida, 2008-2018

|  | 2008 | 2009 | 2010 | 2011 | 2012 | 2013 | 2014 | 2015 | 2016 | 2017 | 2018 |
| --- | --- | --- | --- | --- | --- | --- | --- | --- | --- | --- | --- |
| Sample Size | n=4,710 | n=8,697 | n=11,172 | n=13,268 | n=14,910 | n=16,823 | n=18,369 | n=20,522 | n=21,437 | n=20,676 | n=21,540 |
| Age, mean ±SD | 73.4 ± 5.3 | 73.6 ± 5.4 | 73.9 ± 5.6 | 74.3 ± 5.7 | 74.8 ± 5.8 | 75.2 ± 5.9 | 75.6 ± 6.0 | 75.7 ± 6.1 | 76.3 ± 6.2 | 76.9 ± 6.1 | 77.1 ± 6.1 |
| Female, % | 57.6 | 57.4 | 56.6 | 56.5 | 56.3 | 56.4 | 56.4 | 56.0 | 55.7 | 55.7 | 55.8 |
| Hispanic, % | 99.0 | 99.0 | 98.9 | 98.8 | 98.7 | 98.5 | 98.4 | 98.0 | 97.9 | 97.9 | 97.8 |
| DM, % | 35.5 | 36.9 | 39.5 | 41.4 | 42.3 | 43.8 | 45.1 | 46.5 | 47.3 | 48.9 | 48.6 |
| BP, mean ±SD | | | | | | | | | | | |
| SBP, mmHg |  |  |  |  |  |  |  |  |  |  |  |
| Average BP | 134.1±11.5 | 133.0±11.4 | 133.2±11.6 | 131.5±10.7 | 130.0±10.1 | 128.8±9.2 | 128.5±9.2 | 128.1±8.7 | 129.2±8.8 | 129.2±8.4 | 129.6±8.6 |
| Last BP | 134.0±16.3 | 131.9±16.7 | 132.7±16.7 | 129.7±15.2 | 128.6±14.4 | 127.3±13.0 | 126.5±12.4 | 128.2±13.5 | 128.7±13.4 | 129.0±12.8 | 129.5±12.7 |
| DBP, mmHg |  |  |  |  |  |  |  |  |  |  |  |
| Average BP | 76.5±5.8 | 75.7±5.4 | 74.7±5.2 | 74.0±5.1 | 73.6±4.9 | 73.3±4.6 | 73.0±4.6 | 72.9±4.6 | 73.1±4.6 | 73.2±4.5 | 73.6±4.5 |
| Last BP | 76.3±8.3 | 74.8±7.9 | 74.4±8.0 | 73.5±7.8 | 73.3±7.6 | 72.9±7.3 | 72.5±7.1 | 72.8±7.4 | 73.1±7.3 | 73.3±7.4 | 73.7±7.3 |

SD, Standard deviation; DM, diabetes mellitus; BP, blood pressure; SBP, systolic blood pressure; DBP, diastolic blood pressure

The proportion of hypertensive patients with controlled BP increased substantially after implementation of the hypertension control program in the health system (**Figure 1**). Before program implementation in 2011, overall annual crude BP control rates based on time-average systolic and diastolic BP were in the low 70% range: 70.6% (2008), 73.5% (2009), and 73.1% (2010). BP control rates increased substantially in the first three years of program implementation: 78.5% (2011), 83.5% (2012), and 88.3% (2013). After that, BP control rates stabilized at a high-level plateau: 89.3% (2014), 91.4% (2015), 89.1% (2016), 90.6% (2017), and 89.7% (2018). Control rates calculated based on a single-time point BP at the last clinic visit displayed an analogous pattern, with the exception of a downward period effect in 2015, and the fact that annual BP control rates based on by snapshot BP were systematically lower than those based on within-person time-average BP.

**Figure 1.**
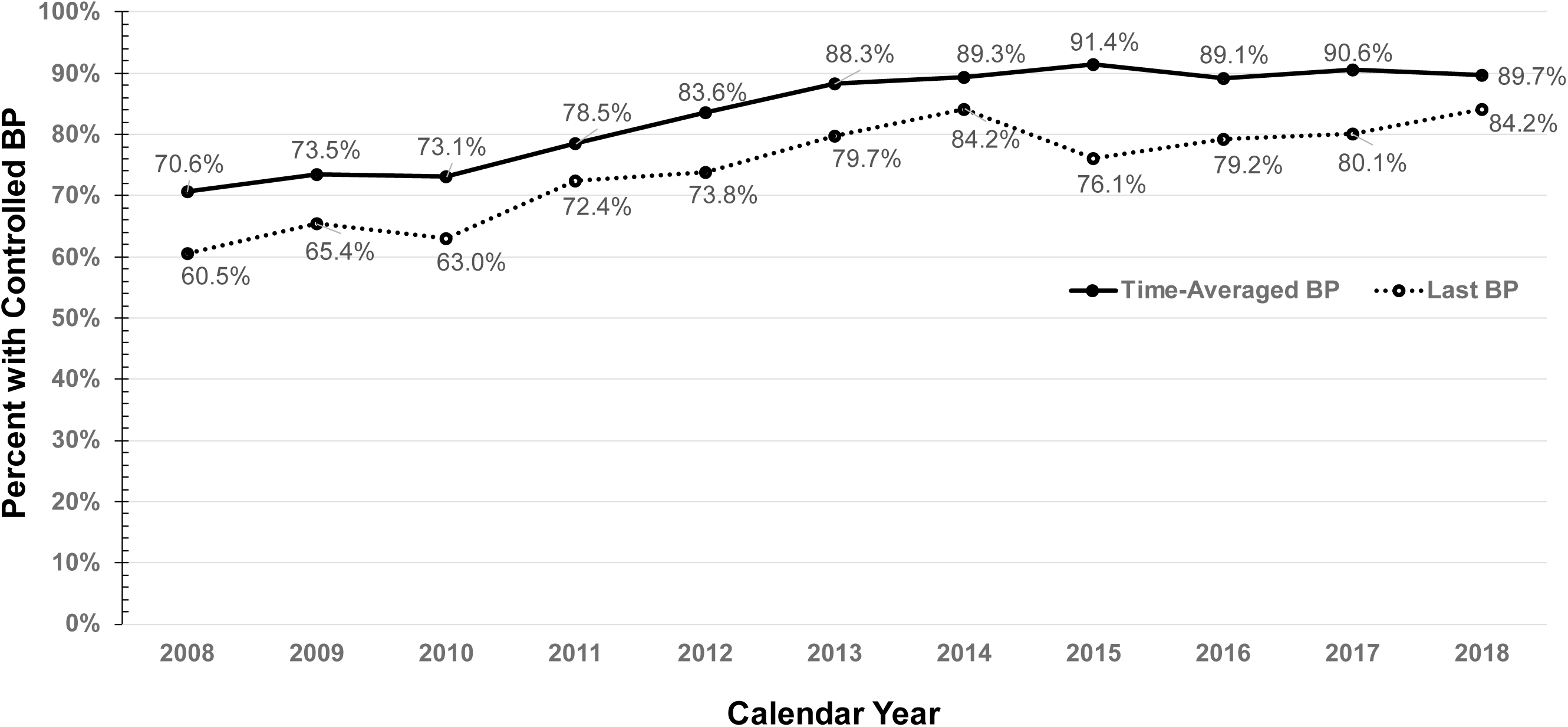
Percentage of patients with hypertension aged 65-89 years old with controlled blood pressure (BP) in a large integrated health system serving Medicare beneficiaries and dually eligible adults in South Florida, 2008-2018. Two unadjusted (i.e., unweighted) BP control metrics are displayed: *time-averaged BP*, defined as the within-person annual average BP of all recorded BP values, and *last BP*, defined based on a single-time point BP; i.e., most recent BP in the year.

Standardized BP control rates for women and men were calculated to account for differences in the distribution of age and diabetes over study period (**Figure 2**). Standardized BP control rates increased in a similar pattern in women and men. Standardized and non-standardized BP control rates also increased in a similar fashion. Standardized BP control rates calculated based on within-person time-average BP were higher than those based on a single-time point snapshot BP in a statistically significant fashion, as indicated by non-overlapping 95% confidence intervals around mean annual BP control rates.

**Figure 2.**
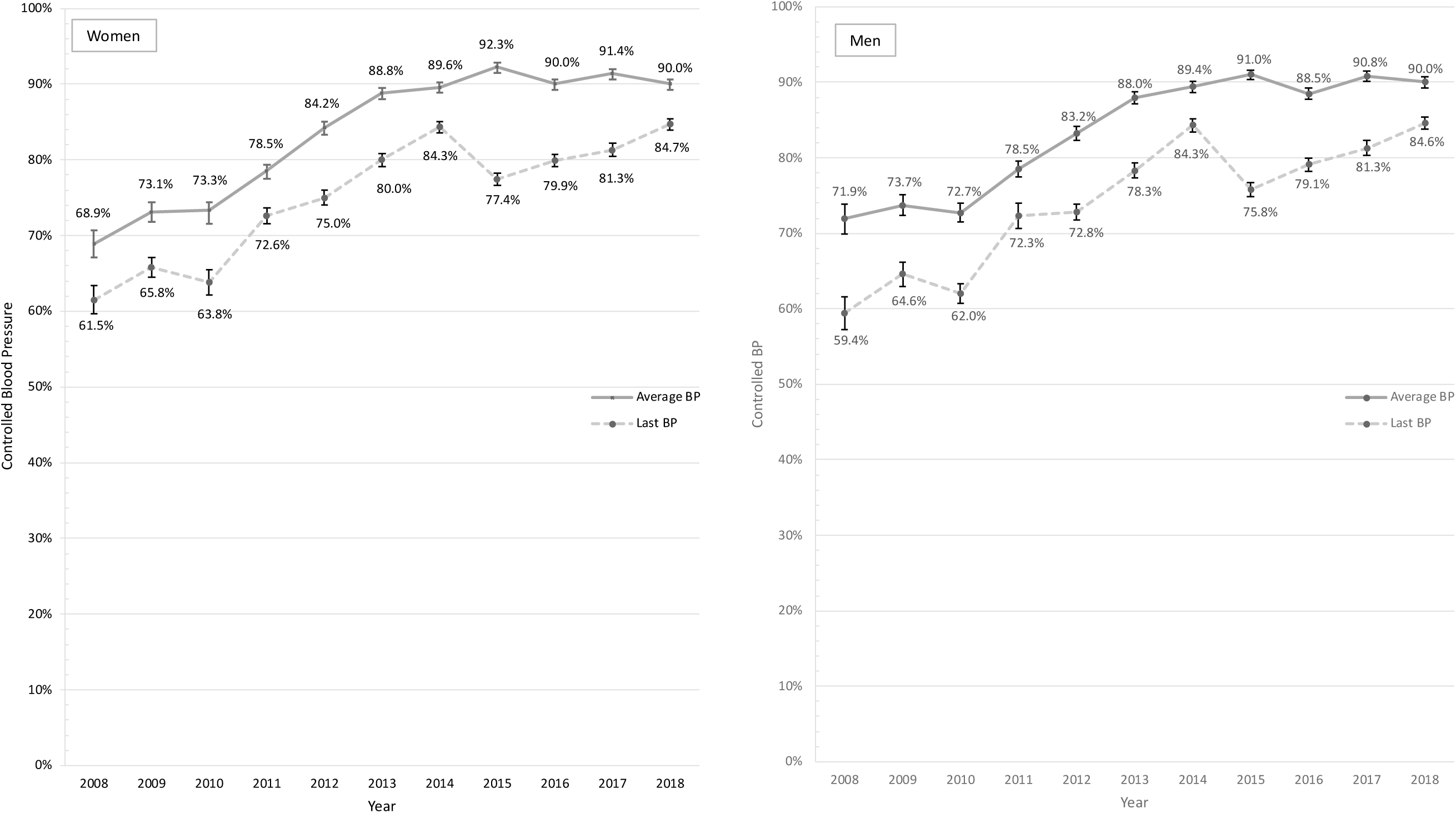
Standardized Annual Blood Pressure Control Rates Adjusting for Differences in Age Distribution and Diabetes Prevalence Over Time. Two standardized blood pressure (BP) control metrics are displayed: one based on within-person annual average BP (time-average BP), and the other based on a single-time point BP; i.e., most recent BP in the year (last BP). Rates were standardized using weights according to the distribution of age and diabetes prevalence in men and women patients in the reference year of 2010. Error bars indicate 95% confidence intervals around each mean BP control rate estimate. Data are from female and male patients aged 65-89 years with hypertension in Leon Medical Centers, Miami, FL, 2008-2018.

Random effects logistic regression models (**Table 2**) compared odds of controlled BP in different hypertension control program implementation phases: before (2008-2010), intermediate (2011-2012), and after fully implemented (2013-2018). In analysis in which BP control was defined based on time-average BP, the overall age-, diabetes-, and gender-adjusted odds of controlled BP based on time-average BP was 5.84 (95%CI: 5.56, 6.14) times higher after program implementation, and 2.14 (95%CI: 2.03, 2.26) times higher in the intermediate phase, as compared to BP control rates before the program. The higher adjusted odds of controlled BP associated with program implementation were similar between women and men. Analysis in which snapshot BP from last visit was used to define BP control yielded results consistent to that estimated based on time-average BP, though strength of associations were not as strong; i.e., OR 1.70 (95%CI: 1.67, 1.77) and OR 2.74 (2.65, 2.84) for odds of controlled BP in intermediate and after implementation phases, respectively, as compared to before-program rates.

**Table 2.** Random effects logistic regression models estimating the adjusted ⱡ odds ratio of controlled blood pressure in female and male hypertensive patients during 3 phases of the hypertension control program ^ŧ^

| BP Control Based on Time-averaged BP |  |  |  |  |  |  |
| --- | --- | --- | --- | --- | --- | --- |
| Characteristic | Overall |  | Female |  | Male |  |
| Program Implementation Periods | OR | (95%CI) | OR | (95%CI) | OR | (95%CI) |
| Pre-Program years (2008-2010) | 1.0 | (Reference) | 1.0 | (Reference) | 1.0 | (Reference) |
| Intermediate years (2011-2012) | 2.14 | (2.03, 2.26)* | 2.17 | (2.03, 2.33)* | 2.10 | (1.94, 2.28)* |
| Post-Program years (2013-2018) | 5.84 | (5.56, 6.14)* | 5.77 | (5.41, 6.16)* | 5.96 | (5.52, 6.44)* |

| BP Control Based on Snapshot BP (Last Available Measurement) |  |  |  |  |  |  |
| --- | --- | --- | --- | --- | --- | --- |
| Characteristic | Overall |  | Female |  | Male |  |
| Program Implementation Periods | OR | (95%CI) | OR | (95%CI) | OR | (95%CI) |
| Pre-Program years (2008-2010) | 1.00 | (Reference) | 1.00 | (Reference) | 1.00 | (Reference) |
| Intermediate years (2011-2012) | 1.70 | (1.63, 1.77)* | 1.68 | (1.60, 1.78)* | 1.73 | (1.62, 1.84)* |
| Post-Program years (2013-2018) | 2.74 | (2.65, 2.84)* | 2.65 | (2.54, 2.78)* | 2.87 | (2.72, 3.02)* |

† OR, odds ratio; CI, confidence interval
‡ Adjusted for age and prevalent diabetes status
\*p-value<0.001

We also examined the concordance with which the two approaches used to calculate BP control, one based on time-average BP and the other on a snapshot of BP, classified similarly subjects as having annual BP controlled or not. Table 3 displays the crude percent agreement in the last 5 years of study for men and women (agreement standardized for differences in the distribution of age and diabetes prevalence across study period was very similar to crude agreement, and thus not shown). In both women and men, percent agreement in the last 5 years of the study ranged approximately from 79% to 84%. Table 3 also displays the kappa statistic, which offers insight into the agreement between approaches in classifying BP control whilst taking into account that some agreement may be expected by chance alone. Kappa statistics in women and men were in the range of.23 to .31 (<.001) and.24 and .33 (<.001), respectively.

**Table 3.** Percent Agreement and Kappa statistic indicating the extent to which two approaches for definition of BP control, one based on within-person annual average BP and the other on the last BP measurement available, agreed in the classification of annual BP control status

| Year | <u>Women</u> |  |  |  | <u>Men</u> |  |  |  |
| --- | --- | --- | --- | --- | --- | --- | --- | --- |
|  | Observed Agreement | Chance Agreement | Kappa | p-value | Observed Agreement | Chance Agreement | Kappa | p-value |
| 2014 | 82.0% | 76.6% | 0.231 | <0.001 | 82.7% | 76.6% | 0.240 | <0.001 |
| 2015 | 78.8% | 71.9% | 0.244 | <0.001 | 78.6% | 71.9% | 0.261 | <0.001 |
| 2016 | 80.7% | 72.8% | 0.288 | <0.001 | 81.1% | 72.8% | 0.307 | <0.001 |
| 2017 | 82.2% | 74.7% | 0.294 | <0.001 | 83.4% | 74.7% | 0.325 | <0.001 |
| 2018 | 83.8% | 76.6% | 0.307 | <0.001 | 84.2% | 76.6% | 0.282 | <0.001 |

## DISCUSSION

This study documented a meaningful, sustained increase in BP control rates up to high levels in older patients with hypertension after implementation of a primary care-based, multicomponent intervention program in a large health care system providing services to Medicare and dual eligible patients primarily of Hispanic origin enrolled in a Medicare Advantage Health Plan in South Florida. Additionally, this study found evidence that annual BP control rates based on last BP reading available in a specific year systematically underestimated those based on the within-person average of BP readings throughout the year.

Our findings are consistent with those that previously demonstrated the positive impact of system-level multicomponent programs on hypertension control ^6–8,22,23^. For example, in 2012, results of a large secondary data analysis study evaluating BP control in patients receiving health care in Veterans Administration medical centers from 2000 to 2010, period in which the Department of Health and Human Services launched the Healthy People 2010 initiative that included a multi-strategy approach to increase BP control, were published ^23^. That study showed steady BP control improvement from 2000 to 2010 in all age, gender, and ethnicity groups studied. The overall percentage of patients with hypertension with controlled BP (<140/90 mmHg) in each month of study period increased from 46.8% in November of 2000 to 76.3% in August of 2010. In 2013, another study reported on the implementation of a large-scale multifaceted quality improvement program in the Kaiser Permanente Northern California integrated healthcare system ^6^. The overall percentage of hypertensive patients classified as having controlled BP (<140/90 mmHg) increased from 43.6% in 2001 to 80.4% in 2009. This study builds on the literature by contributing novel longitudinal data from a more recent calendar time, on an understudied older patient population with a substantially greater proportion of Hispanics and located in a different geographical area than in previous studies, as well as on differences in annual control rates depending on whether they’re based on time-average BP or BP reading at a single-time point (e.g., last visit of the year).

The LMC hypertension control program shared characteristics identified as key in other successful hypertension control program ^3,7^, including: (a) team-based approach; (b) use of standardized protocols for BP measurement and evidence-based guidelines for hypertension management; (c) physician and patient engagement initiatives; and (d) and a robust EHR-based system. The LMC program also included access to in-center pharmacies, a robust transportation system, and an in-house wellness program, which was launched in 2012.

Opened to all patients, whether they had hypertension or not, the latter has offered a broad range of tailored, culturally congruent health promotion activities relevant for cardiovascular health, including group exercise (e.g., Latin dance classes) and individual gym sessions, educational programs on the self-management of chronic diseases, and social integration.

The use of snapshot BP, often the BP reading from the most recent visit, is the traditional operational approach for assessment of the effectiveness of BP control status in health systems. Such a well-established approach facilitates data collection, and the standardization of BP control metrics. Nonetheless, the non-trivial discordance in BP control classification between the approach using last BP reading and annual-average BP, as indicated by a kappa statistic consistent with only fair agreement in this study, along with the fact that longitudinal BP data, which can now be readily gathered by EHR systems, may offer added prognostic insight not captured by snapshot BP ^19,20,24^, it is possible that the use of BP metrics derived from repeated BP readings over time could contribute clinically relevant information for health systems. Further methodological health services research is necessary to determine the potential added value of time-average BP.

Direct comparison of absolute BP control rates observed in this study to those in others require caution. First, our study used real-world BP readings from clinical settings, and included only older adults; in contrast, other studies were conducted in population-based settings and collected BP readings according to rigorous epidemiologic research standards, or in clinical settings that had patients with different age, race/ethnic structures, and/or disease distributions. Second, as compared to snapshot BP readings traditionally used in previous studies, within-individual time-average BP and snapshot BP are inherently different and may lead to relevant differences in BP control status classification as seen in this study. Third, BP control rate calculations in this study had important methodological differences as compared to those calculated according to National Committee for Quality Assurance (NCQA) specifications, including the following: (a) the definition of BP control in this study was kept constant throughout the years – i.e., BP systolic and diastolic less than 140 and 90 mmHg, respectively, regardless of age and disease status – so that changes in BP control rates over time could not be attributed to variations in BP control definition; in contrast, BP was defined as controlled if less than 150×90 mmHg for the majority of those aged 60 years and older as per 2014-2017 NCQA specifications; (b) in this study, we calculated BP control based both on time-average BP, and on most recent visit BP in all hypertensive patients who were aged 65-89 years old and had at least 2 BP measurements (1 or more in each half of the study year), as opposed to rates calculated consistently with audited NCQA specifications based on a substantially smaller sample of patients within the age range of 18-85 years, and that did not required 2 BP readings in the year, and/or that had to follow pre-specified criteria based on when hypertension diagnosis and BP reading were recorded for selection of eligible snapshot BP for rate calculation; and (c) prevalent ESRD, and/or admission to a non-acute inpatient setting during study year were not evaluated for optional exclusion in this study.

This study had several strengths. It focused on an understudied, large minority population of older Hispanics in South Florida, in which the burden of uncontrolled hypertension and other cardiovascular disease-related risk factors is substantial ^14,15^. It leveraged the use of a sophisticated EHRs system, which allowed analysis of BP control rate data changes in a large, clinical setting-based population over a study period of more than 10 years, including 3 years before and 8 years after program implementation.

Study findings should be interpreted in light of important considerations. First, this secondary data analysis study evaluated temporal changes in BP control before and after the implementation of a quality improvement program in a large health system; neither randomization procedures commonly used in pragmatic clinical trials, nor a concurrent control group were in place. Nonetheless, BP control improvement findings and components of the multifaceted program documented here were consistent with those previously reported for programs recognized as successful in BP control. Additionally, medication adherence for hypertension, as per the related NCQA-defined metric calculated as the percent of patients with a prescription for a renin angiotensin system antagonist blood pressure medication who fill their prescriptions often enough to cover 80% or more of the time they are supposed to be taking the medication, improved incrementally and significantly during study period in parallel with BP control improvement from 73.4% in 2010 to 93-95% in the last 3 years of the study (public available performance data reported to CMS in previous years by the health system as part of its administrative routine – not shown here). Thus, our findings are consistent with a direct impact of the hypertension control program. Second, this study focused on the impact of the program as a whole; thus, no inferences about the role of individual program components are appropriate. Third, it should be recognized that program results may vary from one setting to another. This study addressed a culturally congruent program designed for an older adult population of Hispanic origin living in South Florida who were members of a Medicare Advantage plan. Considering that BP control rates may be influenced by a number of factors, including health care access, race/ethnicity, age, and health system resources and management, our results should not be generalized automatically to other population subsets in other geographical locations. It is reassuring, though, that key elements of the studied program here were in line with those included in other programs that led to higher blood pressure control rates implemented in varying populations, which speak to the robustness of health system-and team-based multicomponent programs for BP control ^3,6–8,13,22^. Fourth, this study did not assess the program’s direct impact on adverse distal outcomes of uncontrolled BP; nonetheless, the use of BP control as an intermediate surrogate for prevention of stroke and cardiovascular disease events is well-established.

In summary, this study expands the literature on the control of blood pressure in large populations by contributing data on the successful implementation of a health system-based, multicomponent hypertension control program, which led to sustained high BP control rates in an at-risk, understudied Medicare and dual eligible older patients primarily of Hispanic origin in South Florida. Future research seeking to compare methodologies and metrics are warranted to guide the identification of most feasible, standardized approach that maximizes longitudinal electronic health records data while minimizing data capturing burden in health systems vis-à-vis optimization of the clinical management of BP levels in older adults with hypertension.

## Data Availability

Funder requirements are not applicable, and data will not be shared or will be available upon request.

## ACKNOWLEDGMENTS

Preliminary results from this study were presented at the 2024 Gerontological Society of America Annual Meeting, Seattle, Nov 13-16, 2024. We would like to thank Mr. Adolfo de Varona, Business Intelligence Analyst at Leon Medical Centers, for careful preparation of the analytic dataset for this study. We also would like to thank Dr. Qian-Li Xue for his biostatistical advice and support. Preliminary results were presented at the 2024 Gerontological Society of America Annual Meeting, Seatle, WA. All authors gave their final approval of the version of the article submitted for publication.

